# No support for oxytocin modulation of reward-related brain function in autism: evidence from a randomized controlled trial

**DOI:** 10.1101/2021.03.19.21253900

**Authors:** Annalina V. Mayer, Katrin Preckel, Kristin Ihle, Fabian A. Piecha, Klaus Junghanns, Stefan Reiche, Lena Rademacher, Inge Kamp-Becker, Sanna Stroth, Stefan Roepke, Charlotte Küpper, Veronika Engert, Tania Singer, Philipp Kanske, Frieder M. Paulus, Sören Krach

## Abstract

Autism spectrum disorder (ASD) is characterized by difficulties in social communication and interaction, which have been related to atypical neural processing of rewards, especially in the social domain. Since intranasal oxytocin has been shown to modulate activation of the brain’s reward circuit, oxytocin could be a useful tool to ameliorate the processing of social rewards in ASD and thus improve social difficulties. In this randomized, double-blind, placebo-controlled, crossover fMRI study, we examined effects of a 24 IU dose of intranasal oxytocin on reward-related brain function in 37 men with an ASD diagnosis and 37 age- and IQ-matched control participants. Participants performed an incentive delay task that allows the investigation of neural activity associated with the anticipation and receipt of monetary and social rewards. Apart from a specific interaction effect in a single voxel within the left amygdala during the receipt of rewards, oxytocin did not influence neural processes related to the anticipation or consumption of social or monetary rewards in either group. Exploratory analyses suggested that oxytocin may increase ventral striatum sensitivity to monetary, but not social rewards, in individuals with high levels of self-reported anxiety, depression, alexithymia, and autistic traits irrespective of an ASD diagnosis. There were no significant differences in reward-related brain function between the two groups under placebo. Overall, our results do not support the hypothesis that intranasal oxytocin generally enhances activation of reward-related neural circuits in men with and without ASD without intellectual impairment. How and if oxytocin can be beneficial in the treatment of social difficulties in ASD needs to be addressed by examining moderating influences of individual differences and context on reward-related oxytocin effects.

## Introduction

Difficulties in social communication and interaction are at the core of autism spectrum disorder (ASD). For example, children with autism show low levels of eye contact [1], rarely point at objects to initiate joint attention [2,3], and may struggle to build and maintain friendships [4]. It has been hypothesized that these difficulties relate to a diminished sensitivity to social rewards, such as smiles, praise, or gestures of approval, which leads to a lack of orienting toward social stimuli [5–7]. While behavioral and psychophysiological studies seem to support this notion and often highlight diminished responsiveness particularly to social rewards in individuals with ASD [8–12], neuroimaging studies indicate reduced responses of reward-related brain circuits to both social and non-social rewards (e.g., money), compared to individuals without an ASD diagnosis [13,14]. Specifically, meta-analytic evidence shows robust hypoactivation in the caudate nucleus, the nucleus accumbens located in the ventral striatum, and the anterior cingulate cortex across a variety of reward tasks [14]. These regions are considered key structures within the “reward circuit” [15–17]. This complex circuit consists of several striatal, cortical and midbrain regions, as well as amygdala, hippocampus and specific brainstem structures, and is thought to be a central component underlying the development and control of motivated behaviors [15– 17].

In the last fifteen years, the hormone and neuropeptide oxytocin has gained attention in the fields of psychiatry and social neuroscience as a modulator of social cognition and behavior [18–22]. Importantly, studies in mice have demonstrated that oxytocin plays a crucial role in the processing of social rewards through coordinated activity with serotonin in the nucleus accumbens [23] and by regulating dopamine release in midbrain structures during social interactions [24,25]. In humans, results from functional brain imaging studies suggest that intranasally administered oxytocin can influence neural activity of regions within the reward circuit. For example, oxytocin administration has been shown to increase neural responses to social [26] as well as non-social rewards in the midbrain [27], anterior cingulate cortex and caudate nucleus [28], and to increase perfusion of the nucleus accumbens while viewing pictures of faces [29]. Further evidence for a role of oxytocin in reward processing is provided by investigations of oxytocin receptor distribution in the human brain, showing that oxytocin receptors are located in regions of the reward circuit such as the amygdala, anterior cingulate cortex and brainstem areas, among others [30]. Moreover, a recent study demonstrated that genes involved in the oxytocin pathway are highly expressed in regions that are associated with reward, motivation and learning [31].

With oxytocin being involved in a range of social behaviors [32], it has been suggested that variation in the oxytocin system contributes to the etiology of ASD [33]. A growing body of literature indicates associations of blood oxytocin levels [34–39] and oxytocin pathway genes with the degree of social difficulties or the diagnosis of ASD per se [40–42]. This evidence has led to a rising interest in intranasal oxytocin as a potential treatment of social symptoms in ASD. Several clinical trials have investigated the effects of a single or repeated doses of oxytocin, to date with overall inconclusive results [43,44]. However, clinical studies using functional magnetic resonance imaging (fMRI) suggest that oxytocin may influence regions within the reward circuit in autistic individuals. For example, it has been shown that oxytocin administration enhanced activity in the striatum and prefrontal cortex areas in children with ASD during judgments of social and non-social images, with significantly greater striatal activity during the social vs. non-social task [45]. Another study demonstrated increased connectivity between the nucleus accumbens and cortical regions of the reward circuit after oxytocin administration during two tasks involving social stimuli [46].

To date, only two imaging studies have examined oxytocin effects on reward processing in ASD more directly and have yielded contradictory results. One study testing men with ASD without intellectual impairment reported that intranasal oxytocin specifically increased learning from *social* cues and reinforcement, which was accompanied by a stronger association of reward prediction errors and nucleus accumbens activity compared to placebo [47]. In contrast, another study in children with ASD without intellectual impairment found enhanced activity within regions of the reward circuit only during the anticipation of *non-social* rewards after oxytocin administration [48]. Overall, little is known about the influence of oxytocin on the processing of social vs. non-social rewards in individuals with ASD. Important questions such as whether this potential influence is similar or different in individuals without ASD, and which experimental (e.g., reinforcement learning vs. reward anticipation) and individual factors (e.g., personality, degree of social difficulties, age) might contribute to its efficacy, remain to be answered.

To address these open questions, we conducted a randomized, double-blind, placebo-controlled, crossover fMRI study examining the effect of a single dose of oxytocin on reward-related brain function in 37 men with an ASD diagnosis without intellectual impairment and 37 age- and IQ-matched control participants. Participants performed a well-established incentive delay task that allows an investigation of neural activity associated with the anticipation and receipt of monetary and social rewards [49–51]. The incentive delay task was embedded in a multi-center trial with the overall goal to compare oxytocin effects across different facets of social cognition and affect [52,53]. We hypothesized that participants with ASD, compared to control participants without ASD, would show less pronounced activity within regions of the reward circuit under placebo, specifically during social reward anticipation and consumption. Further, we assumed that oxytocin would trigger an increase in reward-related brain function in ASD, accompanied by faster response times during the anticipation of especially social rewards. The assessed measures of reward sensitivity were consequently expected to be comparable to those of the control group under placebo.

## Methods and Materials

### Participants

Participants were enrolled at the University of Lübeck/University Hospital Schleswig-Holstein in Lübeck and the Max-Planck Institute for Human Cognitive and Brain Sciences in Leipzig between December 2016 and January 2019 (*n*_Lübeck_ = 24, *n*_Leipzig_ = 50). The sample consisted of 37 men with a confirmed ICD-10 diagnosis of Asperger’s syndrome (*n* = 28), infantile autistic disorder (*n* = 6), or atypical autism, (*n* = 3) and 37 healthy male control participants that were matched one-on-one for age (±7 years) and full-scale IQ (±7 IQ-points on the Wechsler Adult Intelligence Scale [54], see Table 1). Eligible participants were German native speakers between 19 and 40 years of age. We excluded participants with an IQ lower than 70, a BMI less than 18 or over 30, those with a frequent use of drugs, alcohol and nicotine (more than 15 cigarettes per day), those with current suicidal tendencies, contraindications to oxytocin (e.g. cardiac arrhythmia), metal implants or any other MRI contraindications, as well as participants who were not able to give consent. Participants receiving concurrent antidepressant (*n* = 2) or other medication (*n* = 2) were required to keep the dosage constant during participation. Participants with ASD were recruited from five university centers offering autism specific diagnosis and counselling across Germany, as well as through online advertisements and leaflets. In the specialized university centers, participants with ASD had undergone standardized diagnostic procedures including the Autism Diagnostic Observation Schedule (ADOS [55]) and the Autism Diagnostic Interview-Revised if parental informants were available (ADI-R [56]; caregivers were available in 70.3% of all cases). Participants with ASD who were not referred directly from an autism specific university center were required to provide a report detailing the diagnostic process of their ASD diagnosis (i.e., whether the diagnosis was based on gold standard diagnostic procedures including ADI-R and ADOS) and a confirmation of their ASD diagnosis based on ICD-10 criteria. Control participants were recruited through in-house databases and from the general population via public notices and online advertisements. Additional exclusion criteria for control participants included a history of neurological, endocrinological or psychiatric disorders based on self-report, current psychotherapeutic or psychiatric treatment, current use of psychotropic medication, first-or second-degree relatives with autism spectrum disorders, and a score of 32 or higher on the Autism-Spectrum Quotient (AQ) [57]. This study was approved by the Ethics Committee of the University of Lübeck, Germany (AZ 15-337) and the German Federal Institute for Drugs and Medical Devices (Bundesinstitut für Arzneimittel und Medizinprodukte; BfArM; Gz: 61-3910-4041063) in Bonn, Germany, and was carried out in compliance with the Declaration of Helsinki of 1975, as revised in 2008. Full informed written consent was obtained from all participants.

**Table 1:**
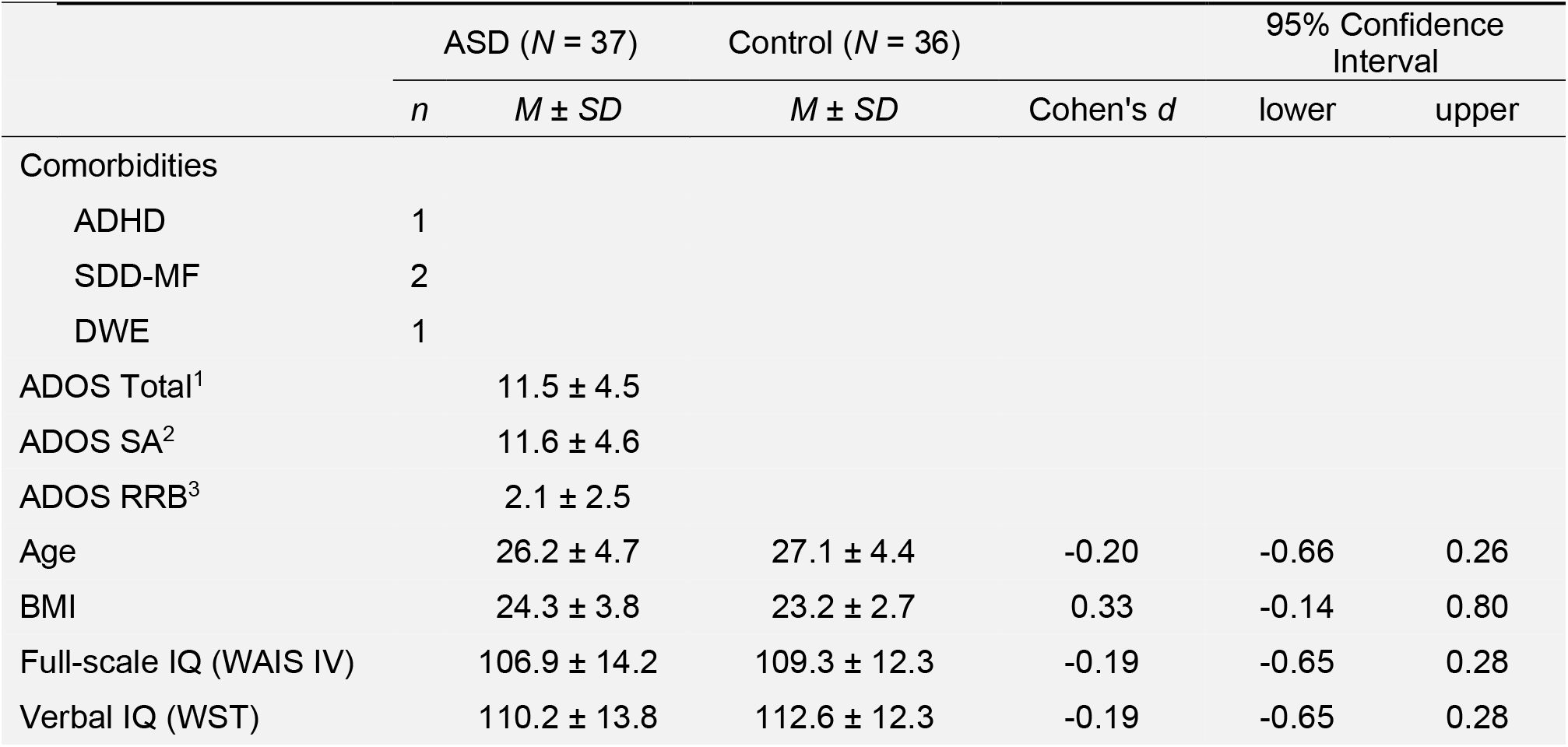
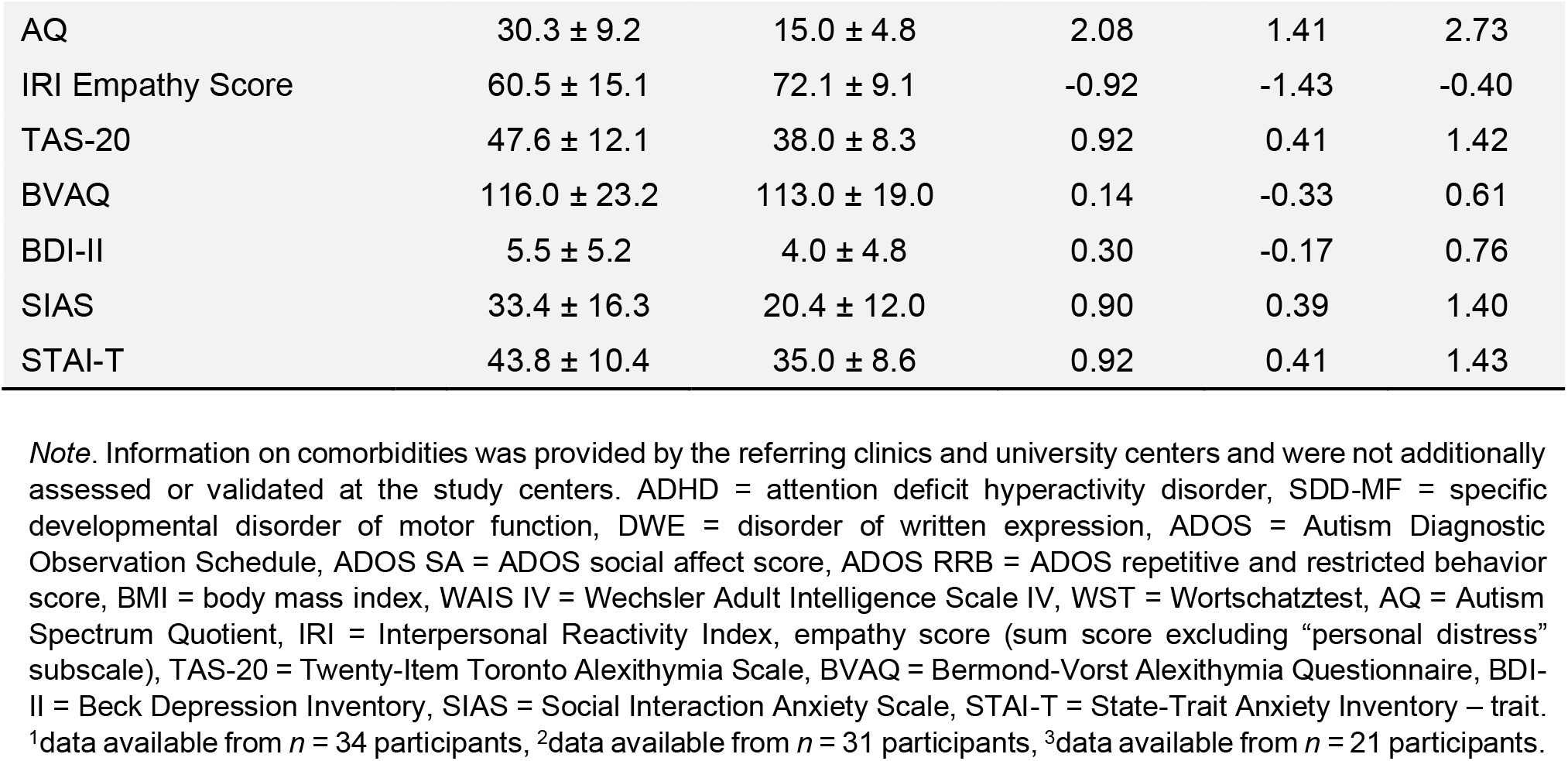
Sample characteristics.

| | ASD ($N = 37$) | | Control ($N = 36$) | | 95% Confidence Interval | |
| --- | --- | --- | --- | --- | --- | --- |
| | $n$ | $M \pm SD$ | $M \pm SD$ | Cohen's $d$ | lower | upper |
| Comorbidities |  |  |  |  |  |  |
| ADHD | 1 |  |  |  |  |  |
| SDD-MF | 2 |  |  |  |  |  |
| DWE | 1 |  |  |  |  |  |
| ADOS Total<sup>1</sup> | | 11.5 ± 4.5 | | | | |
| ADOS SA<sup>2</sup> | | 11.6 ± 4.6 | | | | |
| ADOS RRB<sup>3</sup> | | 2.1 ± 2.5 | | | | |
| Age | | 26.2 ± 4.7 | 27.1 ± 4.4 | -0.20 | -0.66 | 0.26 |
| BMI | | 24.3 ± 3.8 | 23.2 ± 2.7 | 0.33 | -0.14 | 0.80 |
| Full-scale IQ (WAIS IV) | | 106.9 ± 14.2 | 109.3 ± 12.3 | -0.19 | -0.65 | 0.28 |
| Verbal IQ (WST) | | 110.2 ± 13.8 | 112.6 ± 12.3 | -0.19 | -0.65 | 0.28 |

|  |  |  |  |  |  |
| --- | --- | --- | --- | --- | --- |
| AQ | 30.3 ± 9.2 | 15.0 ± 4.8 | 2.08 | 1.41 | 2.73 |
| IRI Empathy Score | 60.5 ± 15.1 | 72.1 ± 9.1 | -0.92 | -1.43 | -0.40 |
| TAS-20 | 47.6 ± 12.1 | 38.0 ± 8.3 | 0.92 | 0.41 | 1.42 |
| BVAQ | 116.0 ± 23.2 | 113.0 ± 19.0 | 0.14 | -0.33 | 0.61 |
| BDI-II | 5.5 ± 5.2 | 4.0 ± 4.8 | 0.30 | -0.17 | 0.76 |
| SIAS | 33.4 ± 16.3 | 20.4 ± 12.0 | 0.90 | 0.39 | 1.40 |
| STAI-T | 43.8 ± 10.4 | 35.0 ± 8.6 | 0.92 | 0.41 | 1.43 |

*Note.* Information on comorbidities was provided by the referring clinics and university centers and were not additionally assessed or validated at the study centers. ADHD = attention deficit hyperactivity disorder, SDD-MF = specific developmental disorder of motor function, DWE = disorder of written expression, ADOS = Autism Diagnostic Observation Schedule, ADOS SA = ADOS social affect score, ADOS RRB = ADOS repetitive and restricted behavior score, BMI = body mass index, WAIS IV = Wechsler Adult Intelligence Scale IV, WST = Wortschatztest, AQ = Autism Spectrum Quotient, IRI = Interpersonal Reactivity Index, empathy score (sum score excluding “personal distress” subscale), TAS-20 = Twenty-Item Toronto Alexithymia Scale, BVAQ = Bermond-Vorst Alexithymia Questionnaire, BDI-II = Beck Depression Inventory, SIAS = Social Interaction Anxiety Scale, STAI-T = State-Trait Anxiety Inventory – trait. <sup>1</sup>data available from $n = 34$ participants, <sup>2</sup>data available from $n = 31$ participants, <sup>3</sup>data available from $n = 21$ participants.

Details on a priori sample size calculations are given in our published study protocol [52]. Briefly, based on meta-analytic effect size estimates available at the time of study planning, we assumed relatively strong effects of oxytocin treatment on neural network activity (*d* = 0.65). A sample of *n* = 88 participants with ASD (and *n* = 88 control participants) would be necessary to reach a desired power of 80% at a significance level of α = 0.001 for two-tailed t-test comparisons of oxytocin effects in a 2×2 cross-over design. Because of difficulties with patient recruitment and resulting significant time delays, the study had to be ended prematurely. Of the enrolled *n* = 74 participants, one control participant discontinued the study after the first MRI session, leaving data from *n* = 73 participants to be analyzed (supplementary Figures S1 and S2).

### Trial Design

This clinical trial was registered in the German Clinical Trial Register (registration number: DRKS00010053) on the 8th of April 2016.

This multi-center clinical trial used a randomized, double-blind, placebo-controlled, cross-over protocol. After an initial screening procedure, eligible participants were invited to three visits at the respective study site. During the first visit, all participants received detailed medical information on intranasal oxytocin and the MRI assessment, after which informed written consent was obtained. Participants with ASD were then randomly allocated to a treatment arm determining the order of treatment (oxytocin first/placebo first). Details on randomization and blinding are given in supplementary methods. Control participants were assigned to the same treatment arms as the participants they were matched to. During the second and third visit, participants received a single dose of oxytocin or placebo intranasally and took part in three independent experiments in the MRI scanner starting approximately 40 minutes after substance administration. Except for the administered nasal spray, the second and third visit included identical procedures and measurements. The visits were 14 days apart to provide a sufficiently long wash-out period.

### Procedure

Interested participants were first screened for main inclusion criteria. For interested control participants, this screening consisted of a short telephone interview, as well as the completion of the Wechsler Adult Intelligence Scale (WAIS-IV) [54] at the respective study site. If a control participant was matched with an already randomized participant with ASD, he was invited to the first study visit. For interested participants with ASD, successful telephone screening was directly followed by the first study visit. In preparation for the first visit, participants received a copy of the informed consent to read. During the first visit, a study physician gave detailed information on risks and benefits of the study and answered the participant’s questions. Following written consent, a study nurse ensured that all inclusion and no exclusion criteria were met. For this purpose, heart rate and blood pressure were measured, and MRI compatibility was assessed using a questionnaire. A brief standardized interview was conducted to exclude possible alcohol abuse or dependence. Subsequently, patients and control participants filled out several health and personality related questionnaires. These included the Toronto Alexithymia Scale (TAS 20) [58,59], the Beck Depression Inventory (BDI-II) [60,61] and the Autism Spectrum Quotient (AQ) [57]. Further, the WAIS-IV [54] was administered to participants with ASD to obtain a valid estimate of full-scale IQ. All participants were instructed to abstain from food, caffeine, and excessive amounts of water 2 hours before the beginning of the next two study visits.

At the beginning of the second and third visits it was ensured that none of the inclusion or exclusion criteria had changed since the last visit. To monitor potential cardiovascular effects of oxytocin, heart rate and blood pressure were measured several times during these visits, starting with two baseline measurements at the beginning of each session. Next, participants were asked to provide a saliva sample for cross-sectional (epi)genetic analyses conducted within the multi-center consortium on ASD [53]. Subsequently, under the guidance of an investigator, participants self-administered 12 puffs of the nasal spray (6 puffs per nostril, each containing 2 IU) over the course of several minutes. A third and fourth measurement of blood pressure and heart rate followed nasal spray administration. Further, current feelings of anxiety were assessed using the state scale of the State Trait Anxiety Inventory (STAI-S) [62]. Approximately 40 minutes after substance administration, participants entered the scanner and performed three independent experiments, which took about 90 minutes. The reward experiment was one of these three experiments. Results from the other two tasks, which are described in our published study protocol [52], will be reported separately. Details on the average latency between substance administration and start of the reward experiment are given in supplementary Table S1. Immediately after the MRI session, blood pressure and heart rate were measured twice, and participants once again indicated their current state of anxiety using the STAI-S. At the end of each session, participants were asked to guess whether they had received oxytocin or placebo. In total, the second and third visits lasted about 2.5 to 3 hours.

To assess additional relevant health and personality measures, participants were asked to fill out several online questionnaires between the second and third visit. These included, among others, the Interpersonal Reactivity Index (IRI) [63,64], the Social Interaction Anxiety Scale (SIAS) [65], the State-Trait Anxiety Inventory – trait (STAI-T) [62] and the Bermond-Vorst Alexithymia Questionnaire (BVAQ) [66]. A complete list of all questionnaires that were assessed but not used in the current data analysis is provided in supplementary methods.

Subjective side effects were assessed by directly asking participants if they had experienced any health-related problems after the second and third visit. Participants were instructed to contact the study team in case of subjectively experienced side effects up to two weeks after substance administration. No side effects were reported.

### Task and Stimuli

The monetary and social incentive delay paradigm (MID/SID) was one of three tasks that participants performed in the scanner. The MID [51] and its adaptation SID [49,50] are well-established paradigms that are used to examine an individual’s neural response to the anticipation and consumption of monetary or social rewards. In the current study, 36 MID and 36 SID trials were mixed and presented in a pseudo-randomized order within a single experiment. During the task, participants were required to press a button whenever a target symbol appeared on the screen. Sufficiently fast responses were followed by a picture of either a face (SID) or a wallet (MID). Blurred pictures were shown when the reaction was too slow (Fig. 1A). Task difficulty was standardized to a hit rate of ∼66% by adjusting the time window for responses to individual response times. A first estimate of individual response times was calculated during a training phase at the beginning of the experiment, and the time window was continuously adjusted to the participant’s performance during the main experiment. In the main experiment, cues preceding the target indicated which type of picture was going to be presented after a sufficiently fast response (Fig. 1B). Circles signaled pictures of wallets and squares signaled pictures of faces. Horizontal lines within the circles and squares further indicated whether or not the picture would contain a reward: three lines signaled a smiling face (social reward) or a wallet filled with coins (monetary reward); a single line signaled a face with a neutral expression or an empty wallet (no reward). For the face stimuli, 26 color photographs displaying 2 different expressions of 13 people (7 female, 6 male) were taken from the NimStim set of Facial Expressions [67]. The money stimuli consisted of 28 self-created color photographs displaying 14 different wallets, each once empty and once filled with coins. Before entering the scanner, participants received detailed instructions on the task and were encouraged to respond as fast as possible to all cue types. A brief test run consisting of 5 trials ensured that the instructions were understood correctly. During the two MRI sessions, parallel versions of the task were used, each with different stimuli.

**Figure 1:**
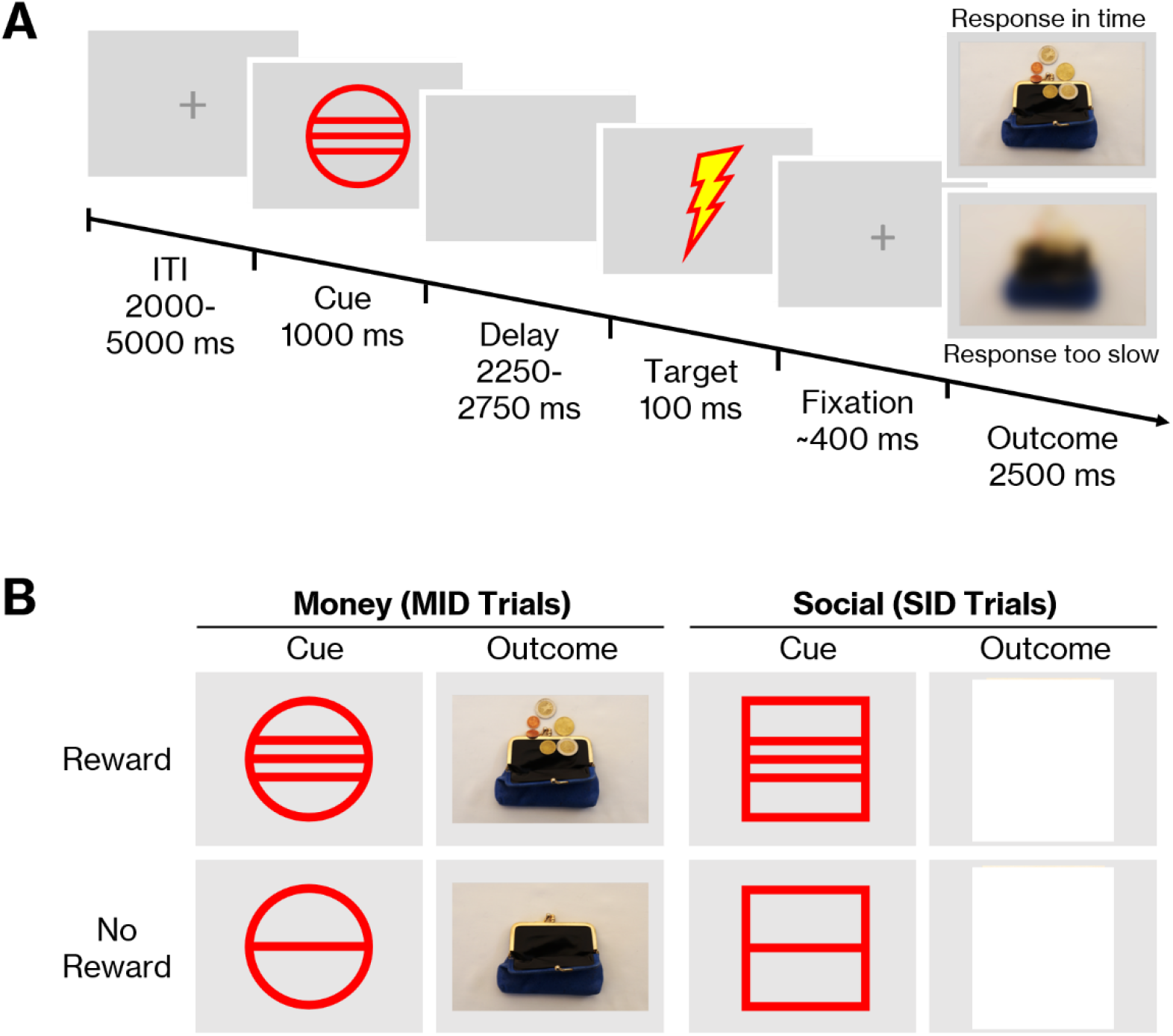
The monetary and social incentive delay paradigm (MID/SID). A: Timing of the task. Participants were asked to hit a button as fast as possible when the target appeared on the screen. Sufficiently fast responses were followed by an image of either a face or a wallet. Blurred images were shown when responses were too slow. B: Social and monetary outcome stimuli and associated cues. To create a reward anticipation phase, a cue indicating the outcome for sufficiently fast responses was presented before the target. Circle cues signaled pictures of wallets, while squares signaled pictures of faces.

**Figure 2:**
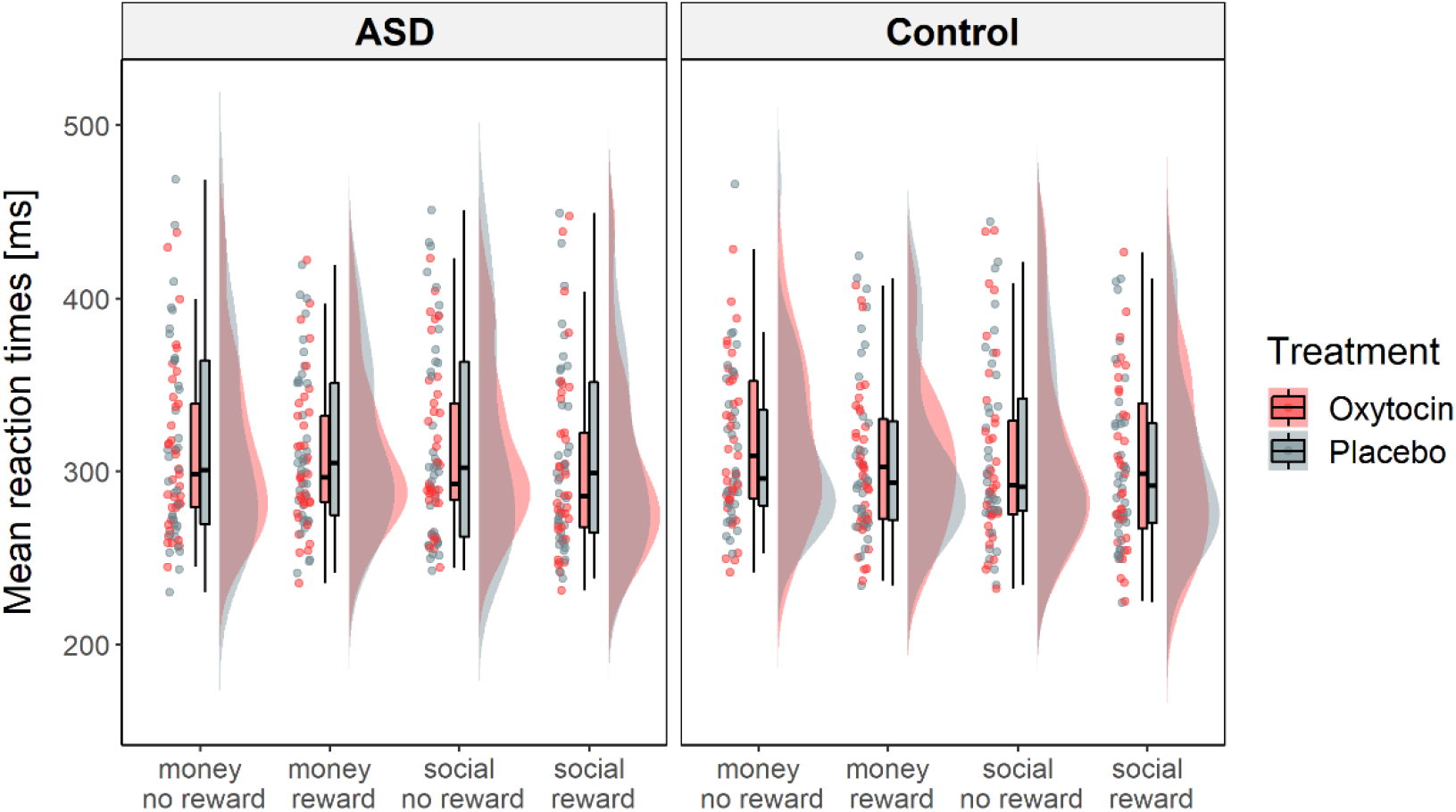
Mean response times for each task type and level of reward intensity. On average, response times were faster when participants anticipated reward compared to no reward. There were no significant differences between the ASD group and control group and no effects of treatment on mean response times.

The experiment was run using the software Presentation® (Neurobehavioral Systems, Inc., San Francisco, CA). Stimuli were presented on a screen positioned behind the MRI that participants viewed through a mirror mounted to the head coil. Participants responded to the target by pressing a button on a fiberoptic response box with the index finger of their right hand. Response times were recorded. At one study site (Leipzig), fMRI assessment was accompanied by eye-tracking and measurements of heart rate, respiration, and skin conductance, which will be analyzed and reported separately. The total experiment, including a brief training phase at the beginning to determine the participant’s average reaction time, lasted about 15 minutes.

### fMRI Data Acquisition

Participants were scanned using a 3T Siemens MAGENTOM Skyra scanner (Siemens, Erlangen, Germany) at the Center of Brain, Behavior, and Metabolism (CBBM) at the University of Lübeck and a 3T Siemens MAGNETOM Skyra fit scanner (Siemens, Erlangen, Germany) at the Max-Planck Institute for Human Cognitive and Brain Sciences in Leipzig. An echo planar imaging (EPI) sequence was used for the acquisition of functional volumes (number of slices = 50, voxel size = 3 × 3 × 3 mm^3^, FoV = 210 × 210 mm^2^, TR = 2000 ms, TE = 27 ms, 90° flip angle). Simultaneous multi-slice imaging (slice acceleration factor = 2) was used for accelerated coverage of the whole brain. A high-resolution anatomical image was acquired for normalization using a T1-weighted 3D MPRAGE sequence (number of slices = 176 (sagittal), voxel size = 1 × 1 × 1 mm^3^, FoV = 240 × 256 mm^2^, TR = 2300 ms, TE Lübeck = 5.49 ms, TE Leipzig = 5.52 ms, 9° flip angle).

### Data Analysis

#### Behavioral Data

Behavioral data including response times and hit rates were analyzed using Jamovi version 1.2.27 [68] and R version 4.0.3 [69]. Trials with response times below 100 ms and above 2000 ms were excluded from the analysis. Mean response times and hit rates were computed for each subject, session, and task condition. Since mean response times were positively skewed, we applied a log transformation [70].

Effects of treatment, group and task conditions on mean response times were examined using a repeated-measures general linear model (GLM). Within-subject factors included task type (money vs. social), reward intensity (reward vs. no reward), and treatment (oxytocin vs. placebo). Group (ASD vs. controls) was entered as a between-subject factor. Further, site (Leipzig vs. Lübeck), treatment arm (oxytocin first vs. placebo first) and order of the incentive delay task within the three sub-experiments (1, 2 or 3) were included in the model as between-subject factors to control for these possible confounds.

In an exploratory approach, we analyzed interactions of treatment and several variables capturing interindividual differences, including age, IQ, anxiety (STAI-T and SIAS sum scores), depression (BDI-II sum score), autistic traits (AQ sum score), empathy (IRI sum score of subscales *empathic concern, perspective taking* and *fantasy*), and alexithymia (TAS-20 and BVAQ sum scores) on mean response times. First, as a measure of individual reward sensitivity, we calculated differences in response times for reward and no reward trials for social and monetary cues, respectively (money: no reward-reward and social: no reward-reward). This was done separately for the oxytocin and placebo sessions. Next, all variables of interest were correlated with behavioral reward sensitivity under oxytocin and placebo using Spearman’s rho as a non-parametric measure of rank correlation. For each variable of interest, the resulting two correlation coefficients were then compared using the approach for dependent overlapping correlations proposed by Steiger [71] as implemented in the cocor R package [72].

To examine possible anxiolytic effects of oxytocin, we calculated a repeated-measures GLM with state anxiety (measured with STAI-S) as the dependent variable. Treatment and time (pre-scan vs. post-scan) were entered as within-subject factors, and group, site, and treatment arm were entered as between-subject factors.

A binomial test was used to determine whether participants were able to identify above chance level which nasal spray they had received. Since some participants indicated the same treatment at both sessions, we compared the proportion of correct guesses per treatment across both sessions to 0.5, instead of analyzing correct guesses per session. For the binomial tests, Bayes factors are provided in addition to the p-values as indicators of evidence for and against the alternative hypothesis.

### Imaging Data

Because of an unexpected loss of fMRI data from one session, one patient was excluded from the analyses. Another patient was excluded due to excessive head motion. fMRI data from the remaining *n* = 35 patients and *n* = 36 control participants were analyzed using SPM12 [73] in MATLAB R2019b [74]. Since the length of the experiment varied according to the subjects’ mean response times, the MRI measurements were terminated manually, which led to variance in the total number of images in the timeseries. The images to be analyzed were defined as all images up to two seconds after the end of the last trial, resulting in 381-383 images per subject and session. The functional volumes were slice-time corrected, spatially realigned, and normalized using the forward deformation fields as obtained from the unified segmentation of the anatomical T1 image. To remove low frequency drifts, the images were high pass filtered at 1/128 Hz.

Statistical analyses were performed on unsmoothed data in a two-level, mixed-effects procedure. The first-level general linear model (GLM) for each subject and session included a total of ten regressors of interest defining the onsets and durations of the four task conditions during the anticipation phase (social:reward, social:no reward, money:reward, money:no reward), the four task conditions during the feedback phase of successful trials (hits) and the onsets and durations of trials of the two task types (money vs. social) during the feedback phase of unsuccessful trials (misses). In addition, the six realignment parameters obtained during preprocessing as well as their first derivatives were included as regressors of no interest to account for noise due to head motion. To further reduce the influence of motion-related artifacts, we used the RobustWLS toolbox [75] to estimate the variance of the noise for each image in the time series and obtain a weighted least squares estimate of the regression parameters. Further, due to slightly different hardware and software configurations during image acquisition at the two study sites, the functional images showed differently distributed luminance values. To prevent signal dropout due to these differences, the implicit masking threshold of the linear models on the first level was lowered to 0.3 (default value in SPM 12: 0.8). Contrast images resulting from the first-level analyses were spatially smoothed with an 8 mm full-width half-maximum isotropic Gaussian kernel before being used in the analyses on the second level.

Several repeated-measures GLMs were implemented on the second level to examine the influence of treatment (oxytocin vs. placebo) and group (ASD vs. controls) on task-related brain activation. For this purpose, we created separate GLMs for each of the following first-level contrast images: main effect of reward intensity (reward > no reward), main effect of task type (social > money), and interaction effect of reward intensity × task type, for both the anticipation and feedback phase (hits only), respectively. Each linear model contained treatment as within-subject factor, and group as between-subject factor. To account for potential variance due to differences in image acquisition, site was entered as covariate. In each model, we also controlled for treatment arm (oxytocin first vs. placebo first) as well as the order of the reward incentive delay task within the three sub-experiments (1, 2 or 3). First, we examined the respective average task effect across groups and treatments. Second, we compared task-related brain activation of the ASD and control group under the placebo condition. Third, we examined effects of oxytocin treatment across both groups. Fourth, we examined interaction effects of group and treatment to assess whether the magnitude of oxytocin effects depended on group.

### Regions of interest and whole-brain analyses

To increase the sensitivity of our analyses, we first examined all effects in a priori defined regions of interest (ROI), which were defined separately for the anticipation and outcome phases. The ventral striatum was chosen as region of interest for the anticipation phase due to its pivotal role in reward processing, especially during the anticipation of rewards [51,76,77]. The bilateral ventral striatum mask consisted of two 8 mm spheres around peak coordinates (MNI coordinates left: −10, 10, −2; right: 12, 14, −4) from a meta-analysis of ventral striatum activation associated with reward anticipation [78]. For the outcome phase, the amygdala was chosen as region of interest because of its associations with reward consumption [49] and intranasal oxytocin effects [79,80]. A bilateral anatomical amygdala mask was created using the automated anatomic labeling atlas (AAL) [81] integrated in the WFU PickAtlas [82] (dilation factor one). All analyses were thresholded at a voxel level of *p* < .05, family-wise error (FWE)-corrected for multiple comparisons within the respective masks.

All contrasts of interest were also explored in the whole brain. A whole-brain mask was used to restrict the analyses to within-brain voxels, since signal from non-brain tissue was sometimes included in the contrast images due to the liberal analysis threshold on the first level. This mask was obtained by calculating average images of normalized grey and white matter tissue maps across all included subjects. The resulting two average images of grey and white matter were summed up to create a single mask. Whole-brain analyses were again thresholded at a voxel level of *p* < .05, family-wise error (FWE)-corrected for multiple comparisons within the whole-brain mask.

### Interactions of treatment and individual difference variables

Similar to the approach used for response times, we explored interactions of treatment and individual differences in age, IQ, anxiety, depression, autistic traits, empathy and alexithymia on reward related brain activation in the regions of interest for the anticipation and outcome phases. For the anticipation phase, we extracted average contrast estimates for money: reward > no reward and social: reward > no reward from the left and right ventral striatum. This was done separately for oxytocin and placebo sessions. Next, all individual difference variables were correlated with average contrast estimates under oxytocin and placebo using Spearman’s rho. Again, for each variable of interest, the resulting two correlation coefficients were then compared using the approach for dependent overlapping correlations proposed by Steiger [71] as implemented in the cocor R package [72]. The same procedure was used for the outcome phase, where contrast estimates were extracted from the left and right amygdala.

## Results

### Behavioral data

Mean hit rates for all task conditions ranged from 63,3% to 66,0% (supplementary Table S2), indicating successful adaptation of task difficulty to subjects’ individual response times. A repeated-measures GLM revealed a significant main effect of reward intensity on log-transformed mean response times (*F*(1,67) = 20.61, *p* < .001, η_p_^2^ = .235), with faster responses during the anticipation of reward (*M* = 307, *SD* = 43.6 ms) compared to no reward (*M* = 315, *SD* = 48.4 ms). There were no significant main effects of group or treatment and no significant interaction effects. Differences in age, IQ, anxiety, depression, autistic traits, empathy, and alexithymia did not significantly moderate treatment effects on reward sensitivity across groups (supplementary Figure S3, supplementary Table S3).

However, treatment influenced state anxiety during the two study visits. There was a significant main effect of treatment (*F*(1,67) = 4.01, *p* = .049, η_p_^2^ = .056) and a significant interaction of treatment and arm (*F*(1,67) = 5.78, *p* = .019, η_p_^2^ = .079). Post-hoc t-tests revealed significant differences in STAI-S sum scores only in the group that received placebo first. Here, anxiety was lower after participants received oxytocin (second session, *M* = 31.9, *SD* = 5.76) as compared to placebo (first session, *M* = 33.5, *SD* = 7.20, *t*(67) = 3.19, *p* = .013, corrected for multiple comparisons using Holm’s procedure [83]). There was no such effect in the group that received oxytocin first (oxytocin: *M* = 33.4, *SD* = 7.57, placebo: *M* = 33.1, *SD* = 8.17, *p* > .05). Further, there was a main effect of time (*F*(1,67) = 5.58, *p* = .021, η_p_^2^ = .077), indicating that participants felt more anxious before MRI scanning than after scanning (pre: *M* = 34.0, *SD* = 6.98; post: *M* = 32.6, *SD* = 7.09). A main effect of group (*F*(1,67) = 8.24, *p* = .005, η_p_^2^ = .110) indicated that the ASD patients felt more anxious than control participants (ASD: *M* = 35.4, *SD* = 7.46; control: *M* = 31.1, *SD* = 4.97).

Participants could not identify above chance level which nasal spray they received: 51.5% of participants correctly identified oxytocin (*p* = .904, Bayes factor_10_ = 0.155) and 53.7% of participants correctly identified placebo nasal spray (*p* = .625, Bayes factor_10_ = 0.182).

### Imaging data

#### Anticipation phase

For the anticipation phase, we first analyzed task related activation of the left and right ventral striatum (Fig. 3). All cues induced significant ventral striatum activation compared to baseline (supplementary Figure S4). However, on average, difference contrasts of interest (social > money, reward > no reward, task × intensity) were not associated with ventral striatum activation. There were no significant differences between patients and control participants in the placebo condition for all contrasts of interest. Further, treatment did not significantly influence task related activation in the ventral striatum and there were no significant group × treatment interactions (Fig. 3). Whole-brain analyses revealed significant average task effects in the occipital cortex and fusiform gyrus for social > money and reward > no reward (supplementary Table S4). There were no significant group differences, effects of treatment, or group × treatment interactions on whole-brain activation.

**Figure 3:**
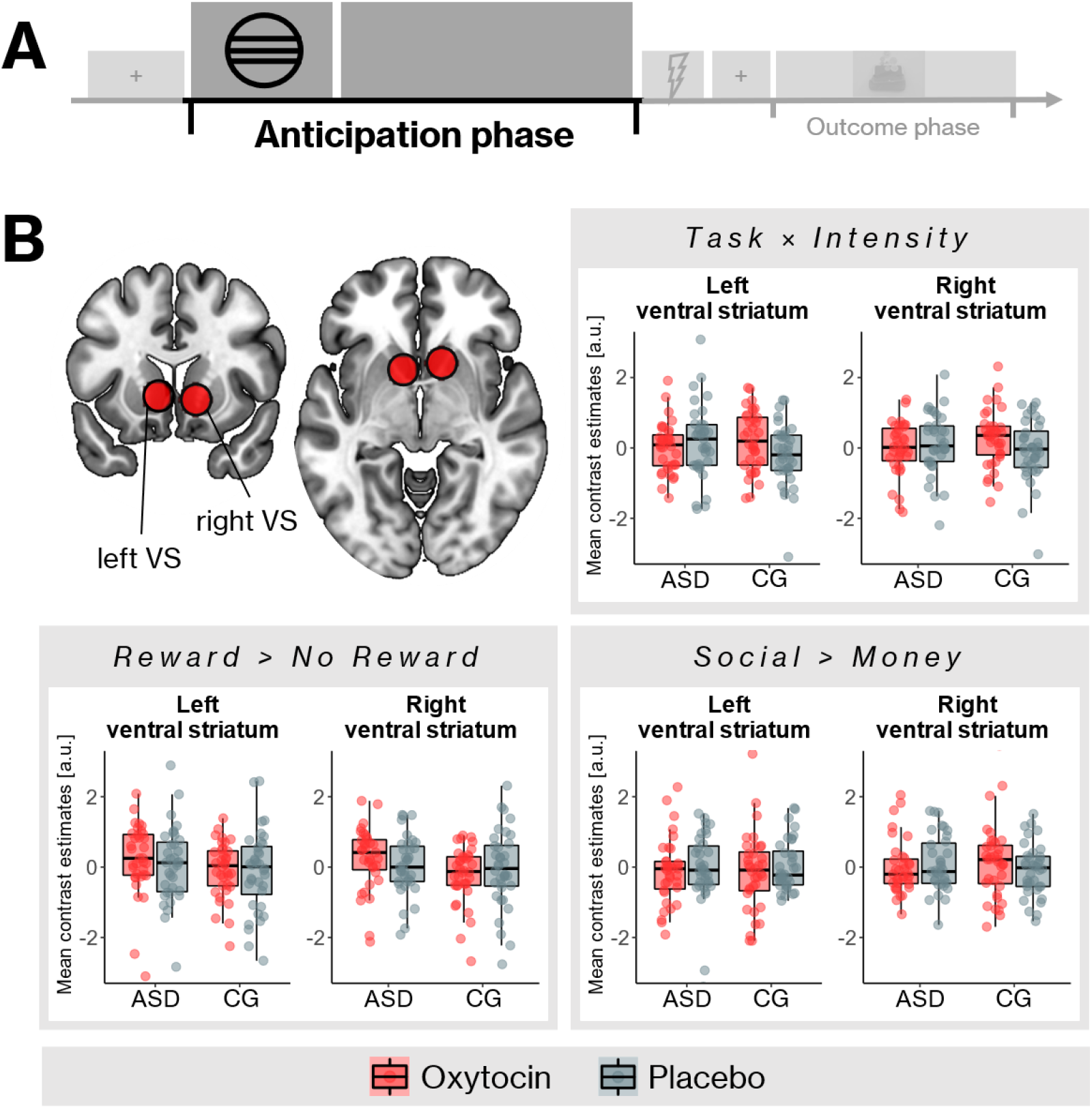
Ventral striatum activation during the reward anticipation phase. A: The anticipation phase was defined as the time interval within each trial between the start of the cue presentation and the target. B: Regions of interest for the ventral striatum (top left) and mean contrast estimates for ASD patients and control participants. Contrast estimates for each contrast of interest were extracted from 8mm spheres around peak coordinates from a meta-analysis examining reward anticipation in the ventral striatum [78]. There were no statistically significant differences between patients and controls, and no significant effects of treatment on task related activation. VS = ventral striatum, CG = control group.

In an exploratory analysis, we examined whether individual differences in age, IQ, mental health, and personality moderated the effects of oxytocin treatment on reward sensitivity in the ventral striatum. Social anxiety (SIAS), autistic traits (AQ), depression (BDI-II) and alexithymia (TAS-20, but not BVAQ) showed significantly different correlations with monetary reward sensitivity after oxytocin compared to placebo (ps < .05, uncorrected; see supplementary Table S5). More precisely, after oxytocin, individuals with higher levels of self-reported social anxiety, depression, alexithymia, and autistic traits experienced higher ventral striatum activation in response to monetary reward cues than individuals scoring low on these traits (ρs = .157 - .383). Under the placebo condition, there was mostly a negative association between these traits and reward sensitivity (ρs = −.205 – −.020). However, none of these traits moderated treatment effects on social reward sensitivity.

#### Outcome phase

For the outcome phase, we first examined task related activation within the left and right amygdala (Fig. 4). On average, the contrasts reward > no reward and social > money were associated with increased activation of the amygdala during hit trials. There were no significant differences between patients and control participants in the placebo condition for all contrasts of interest (Fig. 4). Treatment had a significant effect on the interaction of task and intensity in a single voxel within the left amygdala (MNI coordinates: −21, −4, −25, p(FWE)= .031): compared to placebo, oxytocin enhanced this interaction effect, suggesting that oxytocin led to an increase in differential reward sensitivity for social compared to monetary cues. There were no significant group × treatment interactions on task related amygdala responses. Whole-brain analyses further revealed widespread average task related activation of occipital and superior parietal regions for reward > no reward, occipital and superior parietal regions, orbitofrontal cortex, dorsomedial prefrontal cortex and midbrain structures for social > money, and occipital regions, fusiform gyrus, precuneus and superior parietal regions for the interaction of task and intensity (supplementary Table S6). There were no significant group differences, effects of treatment, or group × treatment interactions on whole-brain activation.

**Figure 4:**
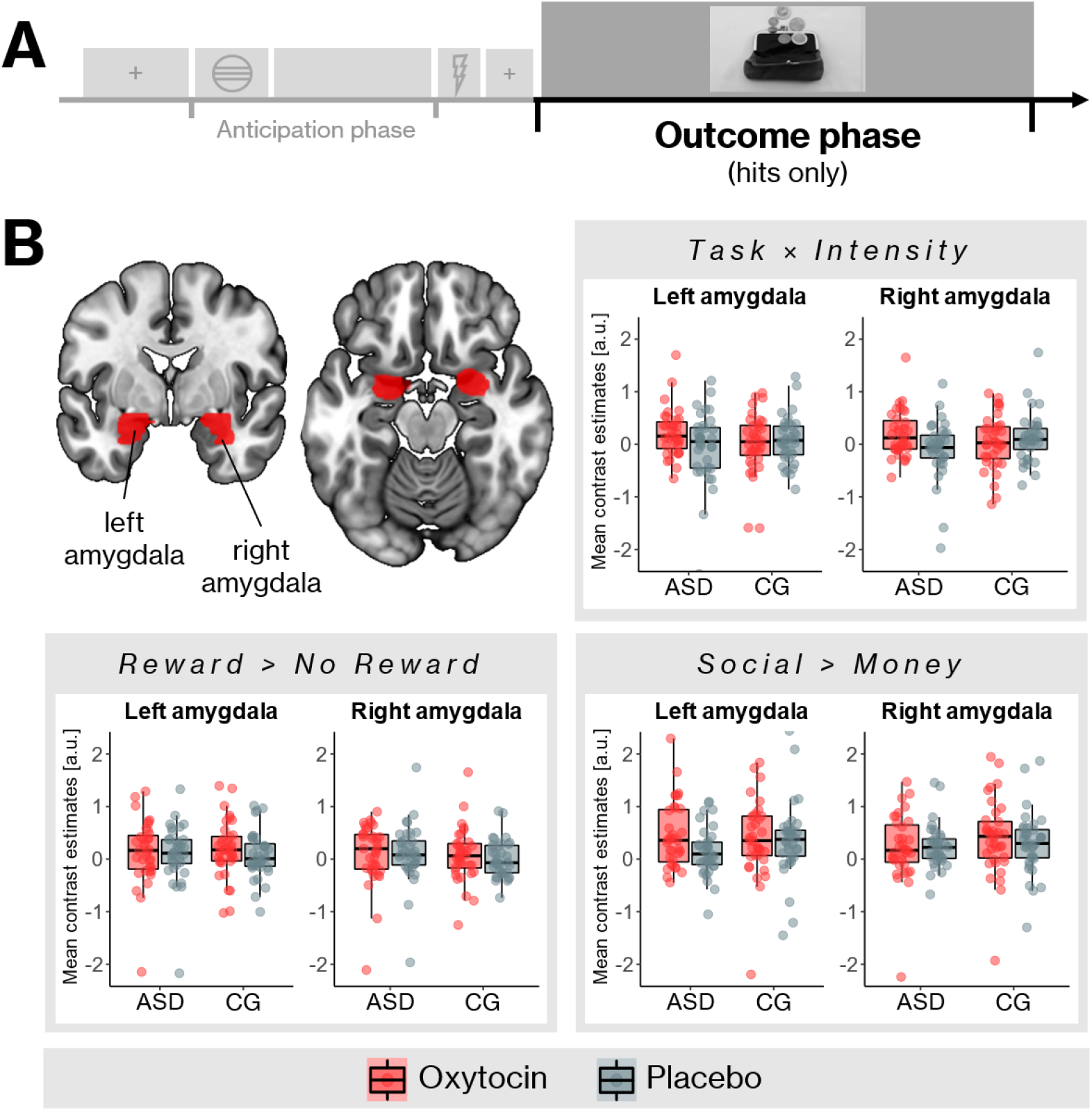
Task related amygdala activation during the outcome phase of hit trials. A: The outcome phase within a trial. We only analyzed hit trials, i.e. trialswith sufficiently fastresponses. B: Regions of interest for the amygdala (top left) and mean contrast estimates for ASD patients and control participants. There were no statistically significant differences between patients and controls. CG = control group.

Again, we explored whether individual differences in age, IQ, mental health, and personality moderated the effects of oxytocin treatment on reward sensitivity in the amygdala. There were no significant differences between correlations of all individual difference variables and reward sensitivity after oxytocin and placebo, suggesting that age, IQ, mental health, and personality did not moderate oxytocin effects on amygdala responsiveness to rewards (supplementary Table S7).

## Discussion

The aim of this randomized controlled trial was to examine acute effects of intranasal oxytocin on the neural correlates of reward processing in male participants with ASD and healthy male control participants. Using a well-established fMRI task, we did not find support for the notion that oxytocin substantially influences brain activation related to reward anticipation or consumption, neither in ASD nor in control participants. Of all analyses across different a-priori specified regions of interest and two task phases, only one contrast yielded a statistically significant effect of oxytocin on brain activation. Further, under placebo there were no statistically significant differences in reward related brain function between subjects with and without ASD. Exploratory analyses suggested that oxytocin might increase ventral striatum sensitivity to monetary rewards in individuals with high levels of self-reported anxiety, depression, alexithymia, and autistic traits.

The only oxytocin effect that could be detected was an enhancement of left amygdala responsiveness to the presentation of social rewards in the outcome phase. More precisely, oxytocin enhanced the interaction of task and reward intensity across both groups in a single amygdala voxel. This means that after oxytocin, this area was more sensitive to differences between neutral and smiling faces, as compared to the difference between pictures of empty and full wallets. This result suggests a relative increase in sensitivity to the reward value of faces, but not an increase of activation in response to faces *per se*. A general oxytocin effect on amygdala activation during face processing has been shown in several studies in adults with and without ASD, which has been interpreted as a neural mechanism underlying oxytocin’s anxiolytic effects [21,79,84–87]. While our findings do not point in the direction of a general enhancement of amygdala activation in response to faces, they can be interpreted as support for the social salience hypothesis of oxytocin, which states that oxytocin modulates orientation to relevant social cues [88]. However, it should be highlighted that this effect was only found within a single voxel, which of course only comprises a small area of the left amygdala. Due to this extremely circumscribed area, and since this effect was the only significant one in a series of comparisons across different task phases and regions, this result should be interpreted with caution.

Overall, the results from the present study are in line with current meta-analytic evidence suggesting that, on average, intranasal oxytocin only has small and sometimes non-significant effects on social cognition and social functioning in individuals with ASD [43,89]. This evidence is further supported by a recent large-scale clinical trial showing that daily administration of oxytocin did not significantly influence social behaviors in 106 men with ASD [90]. However, meta-analyses average across a wide range of tasks and paradigms, which might mask highly context-specific effects of intranasal oxytocin that emerge through general mechanisms such as anxiety reduction [22,91]. Yet, our findings conflict with results from previous neuroimaging studies highlighting the potential of intranasal oxytocin to influence regions within the brain’s reward circuit in individuals with and without ASD [26–29,47,48].

The predominant absence of statistically significant oxytocin effects on brain function could be due to several factors. For example, considering that there is strong evidence linking endogenous oxytocin to reward processing and motivation [23,24,92], one might argue that the method used for administering oxytocin is not ideal. Although some studies suggest that oxytocin is effectively delivered to the brain after intranasal administration [93,94], it has been debated whether the widely used dosage of 24 IU and administration by conventional nasal spray devices are optimal in this regard [95,96]. Possibly, indirect methods of increasing endogenous oxytocin levels, such as the administration of selective melanocortin receptor agonists [97], or the application of oxytocin metabolites [98] could lead to stronger effects than intranasal application. However, the dosage and form of administration used in the current study are identical to those used in previous studies showing an effect of oxytocin on reward processing in individuals with and without ASD [27–29,48]. Moreover, while oxytocin did not influence task-related behavior and brain activation in the current study, we did find a significant anxiolytic effect after oxytocin compared to placebo, indicating that oxytocin indeed reached the brain after administration. It is therefore unlikely that the absence of statistically significant oxytocin effects is solely related to details of the administration protocol.

Another possibility is that intranasal oxytocin effects are tied to specifics of the applied reward task, which could also explain the heterogeneity of the results in oxytocin trials examining reward processing and related brain activation in ASD To date, only two other neuroimaging studies have examined oxytocin effects on reward processing in ASD, and both used conceptually different approaches. While the first study [48] used an incentive delay task, participants of the second study [47] performed a reinforcement learning task. Because of methodological similarities, one might expect the results from the current study to be in line with results from the first study using the incentive delay task. However, even though the monetary or social incentive delay task is one of the most often used tasks to examine reward processing [77], several variations exist that may influence the magnitude of potential oxytocin effects. For example, the choice of stimuli (showing the amount of money won in numbers [48] vs. pictures of money, smiling vs. laughing faces) could influence the perceived value of the rewards and the subjective relevance of the task. Since it has been discussed that oxytocin modulates stimulus processing depending on personal relevance of the stimuli [99], different experimental designs may lead to different results with regards to the oxytocin effects. However, due to the current scarcity of clinical studies investigating oxytocin effects on reward processing in ASD, we can only speculate about the influence of experimental design on the respective outcomes at this point.

Another reason for the predominant absence of general oxytocin effects might be related to an overestimation of effect sizes based on early oxytocin trials. Only a few years ago, oxytocin seemed to be a highly promising treatment option for ASD, with an estimated combined effect size of *d*=0.57 [100]. Subsequent studies examining the effects of intranasal oxytocin on social cognition and behavior have however produced inconsistent results, and some of the earlier studies could not be replicated [101– 103]. Recent research indicates that the median effect size across human oxytocin studies is 0.14, and most studies examining intranasal oxytocin interventions do not have sufficiently large samples to reliably detect effects of this magnitude [104]. Although our sample is considerably larger than the average sample size in oxytocin intervention studies with neurodevelopmental disorders (27.4 participants on average [89]), this suggests that the problem of data insensitivity might also be present in the current sample. However, previous studies on reward-related brain function have shown oxytocin effects even in samples of 15 [47] and 28 [48] individuals with ASD. If these results reflect true effects of oxytocin treatment on reward processing with roughly accurate effect size estimates, we would also expect to see effects in our sample.

While intranasal oxytocin did not have significant effects on reward-related brain function on average, our results suggest that oxytocin efficacy is influenced by individual differences in autistic traits, anxiety, depression, and alexithymia during monetary reward anticipation. After oxytocin administration, individuals with higher levels of self-reported social anxiety, depression, alexithymia, and autistic traits showed higher ventral striatum activation in response to monetary cues than individuals scoring low on these traits. Although this contradicts our hypothesis of oxytocin effectiveness specifically in the social domain, this finding fits well with accounts stating that oxytocin effects depend on personality and other individual factors [91]. Along these lines, previous studies have identified social anxiety [105], alexithymia [106–108] and autistic traits [109,110] as potential moderating factors, which has strengthened the hope that intranasal oxytocin may be specifically effective in individuals with a diagnosis of mental disorder. However, when we examined group × treatment interactions on reward-related brain activation in the present study, there were no significant differences between the control and ASD groups in terms of oxytocin effects. The absence of significant group × treatment interactions is surprising considering that on average, the ASD group showed significantly higher scores for autistic traits, anxiety, depression, and alexithymia than the control group, which is comparable to findings from previous clinical studies [107,108,111]. Since these results are based on an exploratory approach, it will be up to future studies to evaluate the robustness of the modulatory effects of these traits, and to uncover possible reasons for the associations between these social traits and reward sensitivity in a non-social domain.

In addition to the absence of significant group × treatment interactions, we did not find significant differences between the two groups under placebo, neither during the anticipation nor the consumption of social and monetary rewards. This result stands in contrast to a hypothesized social “wanting” dysfunction underlying autistic symptoms [6], which posits atypical reward processing specifically during the anticipation of social rewards. This supposed deficit is at the heart of accounts depicting autism as an “extreme case of diminished social motivation” [112]. However, results from previous neuroimaging studies using a monetary and/or social incentive delay task do not support the notion of a social wanting dysfunction. While two studies showed reward circuit hypoactivation especially during the anticipation and receipt of monetary but not social rewards [113,114], one study showed hypoactivation only during the receipt of social rewards, which corresponds to social “liking” [115]. None of these studies reported pronounced hypoactivation of the reward circuit during social reward anticipation, that is, social “wanting”, in individuals with ASD. These contrary findings are accompanied by criticisms of the social motivation hypothesis, demonstrating that this assumption is contradicted by the testimony of individuals with ASD, and that it often has negative effects on autism research and the development of treatments [116]. Thus, reducing difficulties in social communication and interaction in ASD to disrupted reward-seeking tendencies in social contexts dismisses the inconsistencies within the neuroscientific literature on reward processing, and might be harmful to individuals with an ASD diagnosis.

Our results are also in contrast to meta-analytic evidence showing a broader range of atypical reward processing in ASD [14]. A possible explanation is that atypical reward processing is not universally linked to the pathology of ASD, but rather depends on certain individual factors that cannot be assessed in our relatively homogeneous all-male adult sample. For example, it has been discussed that autistic children and adolescents might show a more pronounced alteration of striatal activation during social reward tasks compared to adults [14], which aligns well with results from studies examining reward processing in adolescents without an ASD diagnosis [117]. Further, more severe autistic symptoms seem to be linked to greater caudate hypoactivation [14]. This could explain the absence of statistically significant differences between our ASD group with relatively mild symptoms and the age- and IQ-matched control group. Taken together, the findings of our study challenge the assumption that atypical reward processing is a primary cause behind social difficulties in ASD. Instead, it seems likely that individual differences in reward processing exist across autistic and non-autistic individuals alike.

## Conclusions

Overall, our results do not support the hypothesis that intranasal oxytocin generally enhances activation of reward-related neural circuits in men with and without ASD. In line with recent evidence, the effects of intranasal oxytocin are likely too subtle to fundamentally shape reward circuitry regardless of context. Instead, oxytocin efficacy may be influenced by features of the experimental design, as well as individual differences in autistic traits, depression, anxiety, and alexithymia irrespective of an ASD diagnosis, especially for non-social rewards. Moreover, our results raise doubts about the hypothesis that reward processing is universally altered in ASD. In light of these findings, the use of oxytocin to improve social reward processing does not appear to be a promising avenue for improving social difficulties in ASD.

## Data Availability

The datasets generated and analyzed during the current study are not publicly available due to them containing information that could compromise research participant privacy and consent. Fully anonymized data are available from the corresponding author on reasonable request.

## Acknowledgements

We would like to thank Johanna Klose, Kerstin Neubauer, Franca Schwesinger, Katharina Ohm, Sarah Rösch and Lisa Nix for their help as student assistants; Janine Baumann, Nora Czekalla, Jöran Lepsien, Toralf Mildner, André Pampel, Manuela Hofmann, Sylvie Neubert, Mandy Jochemko, Anke Kummer, Simone Wipper, Nicole Pampus and Domenica Klank for their assistance in neuroimaging; David Stolz, Laura Müller-Pinzler and Martin Göttlich for their assistance during programming and data analysis; Norbert Brüggemann, Henrike Hanßen, and Anna Kosatschek for their assistance as study physicians; Natalia Chechko, Susanne Stickel (Aachen), Gerti Gerber (Marburg), Daniel Alvarez-Fischer, Veronika Thorns (Lübeck), Stefan Ehrlich, and Veit Rössner (Dresden) for their efforts during participant recruitment; Henrik Grunert and Kerstin Traeger for technical assistance; Matthias Collier, Kathrin Scheibe and Silke Hauer from the Clinical Trial Centre Leipzig who provided data management tools and randomization; and Anne-Kathrin Wermter for her kind assistance during the preparation of this study.

