## Supplementary Material for "No support for oxytocin modulation of reward-related brain function in autism: evidence from a randomized controlled trial"

### SUPPLEMENTARY METHODS

#### *Randomization and Blinding*

Randomization was carried out by the Clinical Trial Center (ZKS) Leipzig independently from the recruitment process and data acquisition. The ZKS created blocked randomization lists separately for each study site with a 1:1 allocation ration and varying block sizes of 2 or 4. According to these lists, participants were allocated to either arm A (oxytocin at first visit, placebo at second visit) or arm B (placebo at first visit, oxytocin at second visit) and received a site-specific medication number. Additionally, the order of experiments during the fMRI sessions was randomized within each treatment arm using blocked randomization with a block size of 6. Blinding was carried out by an independent pharmacist at the Clinic Pharmacy of Heidelberg University Hospital, who received the list of medication numbers and their corresponding treatment arm from the ZKS Leipzig. Oxytocin (Syntocinon® Nasal Spray) and placebo were filled into nasal spray bottles of identical appearance and labeled with a medication number and the according administration sequence (bottle A for first visit, bottle B for second visit). The placebo nasal spray contained all inactive ingredients of the oxytocin nasal spray. Both nasal sprays were identical in appearance and smell. The two bottles were sealed in non-transparent foil, labeled with the respective medication number, and sent to the respective study sites. After receiving a participant's informed written consent, a study nurse filled in an online form provided by the ZKS asserting that all inclusion and no exclusion criteria were met, and subsequently received the medication number and the order of experiments for the respective participant. The center-specific randomization lists were not accessible to investigators engaged in recruitment, data acquisition (including study nurses, study physicians and research assistants), and data analysis. Consequently, all investigators involved in the conduct of the study as well as all participants were blinded to the order of treatment.

*Additional testing and questionnaires*

Over the course of the study, participants were asked to fill out several questionnaires assessing health and personality traits. Since the study was embedded in a Germany-wide research network on mental disorders, some of the data was collected for cross-sectional studies with a focus on understanding mental health an illness in a multidimensional approach [1]. While only some of the questionnaires are used in the current data analysis, all questionnaires and other psychometric tests will also be used for the analyses of the other functional paradigms of this clinical trial. To keep the workload low for participants at the beginning of the study, the assessment battery was divided into two halves and collected at two time points.

The first time point was during the first study visit. The questionnaires and tests collected during the first visit included a questionnaire about basic sociodemographic information, a handedness questionnaire [2], the 53-item Brief Symptom Inventory [3] assessing clinically relevant psychological symptoms, the Toronto Alexithymia Scale (TAS 20) [4,5], the Beck Depression Inventory (BDI-II) [6,7] and the Autism Spectrum Quotient (AQ) [8]. Moreover, participants completed the Trail Making Test A & B [9] as a measure of visual attention and task switching. Verbal intelligence was assessed using a short vocabulary test (WST) [10].

Between the second and third visits, participants filled out the second half of the questionnaires, which were provided online to allow the participants convenient access from their homes. The questionnaires included the WHO Disability Assessment Schedule 2.0 [11], the Childhood Trauma Screener [12], the Behavioral Inhibition and Behavioral Activation Scale [13], the Barratt Impulsiveness Scale (short form BIS-15) [14], the Positive and Negative Affect Schedule [15], the Interpersonal Reactivity Index (IRI) [16,17], the Social Interaction Anxiety Scale (SIAS) [18], the State-Trait Anxiety Inventory – trait (STAI-T) [19], the Cognitive Emotion Regulation Questionnaire [20] and the Bermond-Vorst Alexithymia Questionnaire (BVAQ) [21].

Of all health and personality related questionnaires collected during the clinical trial, only those related to depression (BDI-II), anxiety (SIAS, STAI-T), autistic traits (AQ), empathy (IRI) and alexithymia (TAS-20, BVAQ) were used for the current data analysis.

**SUPPLEMENTARY FIGURES**

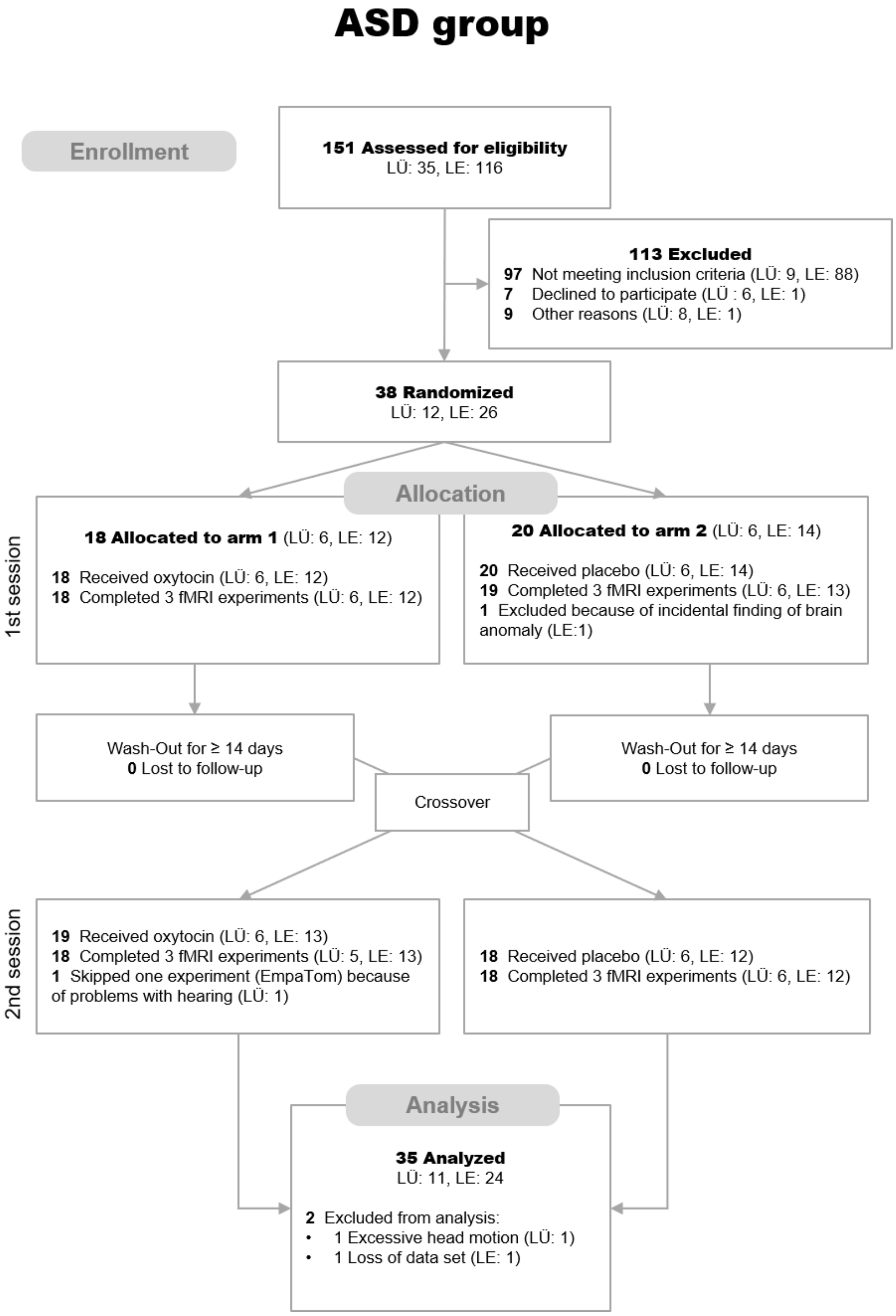


**Figure S1: CONSORT flow diagram for the ASD group.** LÜ = Lübeck, LE = Leipzig. The number of participants indicated in the “Analysis” section refers to those included in the fMRI analyses.

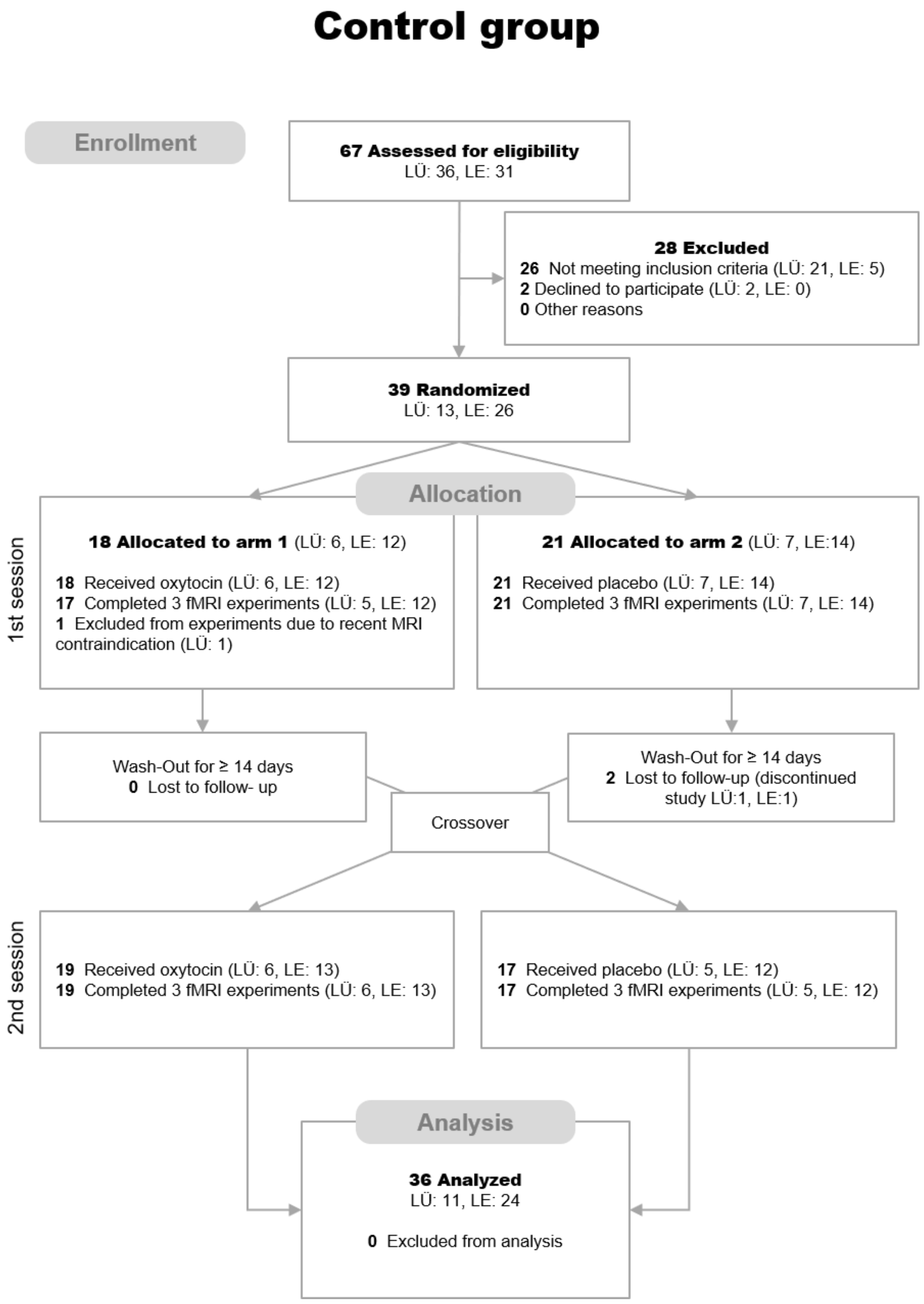


**Figure S2: CONSORT flow diagram for the Control group.** LÜ = Lübeck, LE = Leipzig. The number of participants indicated in the "Analysis" section refers to those included in the fMRI analyses.

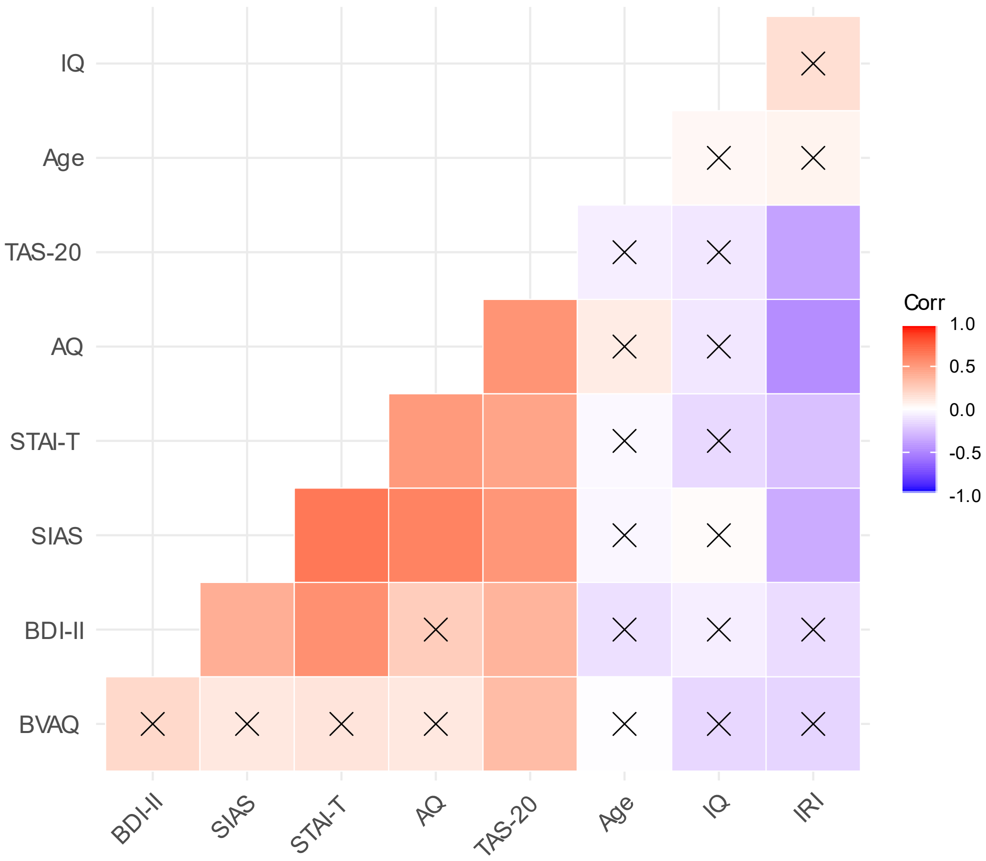


**Figure S3: Intercorrelations of age, IQ, mental health, and personality scores.** Spearman correlation coefficients based on data from *N* = 37 ASD participants and *N* = 36 control participants. Non-significant correlations are marked with an x.

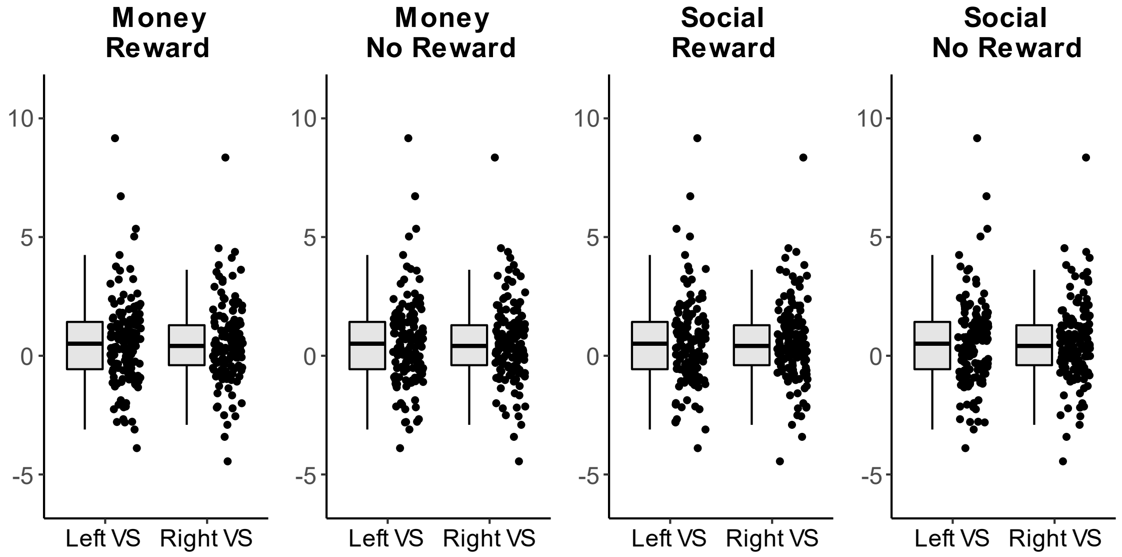


**Figure S4: Contrast estimates for all cues against baseline in bilateral ventral striatum.** All cues induced significant activation of the ventral striatum compared to baseline. $N = 71$.

**SUPPLEMENTARY TABLES**

**Table S1:** Mean latencies between nasal spray administration and start of reward experiment.

| | ASD | | Control | |
|---|---|---|---|---|
| Order | *N* | Latency [min] (*M* ± *SD*) | *N* | Latency [min] (*M* ± *SD*) |
| 1 | 12 | 40.4 ± 6.47 | 12 | 37.8 ± 5.22 |
| 2 | 13 | 62.7 ± 8.93 | 13 | 60.3 ± 10.7 |
| 3 | 12 | 88.1 ± 4.43 | 11 | 87.9 ± 10.3 |

*Note.* The monetary and social incentive delay paradigm (MID/SID) was one of three experiments that participants performed in the MRI. The order of the three experiments was randomized. This led to variance in latencies between nasal spray administration and start of the incentive delay paradigm. There were no statistically significant differences between mean latencies of ASD patients and control participants (two-way ANOVA with factors order and group; main effect group: $p$ = .364).

**Table S2**: Hit rates for each task condition.

| | ASD | | | Control | | |
|---|---|---|---|---|---|---|
| | *M* | Median | *SD* | *M* | Median | *SD* |
| money no reward | 64.6% | 63.9% | 5.1% | 63.3% | 63.9% | 6.0% |
| money reward | 66.0% | 63.9% | 3.8% | 64.8% | 66.7% | 5.4% |
| social no reward | 64.6% | 66.7% | 6.3% | 64.2% | 63.9% | 5.7% |
| social reward | 65.8% | 66.7% | 5.4% | 65.2% | 65.3% | 4.5% |

*Note.* Task difficulty was adapted to each participant's mean response time to achieve a hit rate of ~66% for each task condition. A first estimate of individual response times was calculated during a training phase at the beginning of the experiment, and the time window for responses was continuously adjusted to the participant's performance during the main experiment. $N = 73$.

**Table S3:** Correlations of behavioral reward sensitivity and individual difference variables across all participants after oxytocin and placebo.

| | Money | | | Social | | |
|---|---|---|---|---|---|---|
| | Oxytocin | Placebo | *p*(comparison) | Oxytocin | Placebo | *p*(comparison) |
| Age | .070 | .136 | .654 | -.193 | .074 | .099 |
| IQ | -.045 | .027 | .629 | .041 | -.049 | .579 |
| AQ | -.224 | .006 | .118 | .014 | -.064 | .630 |
| TAS-20 | .039 | .001 | .800 | .062 | -.038 | .537 |
| BVAQ | .060 | **.244** | .209 | .096 | .143 | .768 |
| BDI-II | .187 | **.232** | .757 | .086 | -.044 | .423 |
| IRI | .086 | -.047 | .369 | .205 | .121 | .599 |
| SIAS | -.163 | .081 | .099 | .029 | -.159 | .245 |
| STAI-T | .040 | .087 | .754 | .136 | -.125 | .108 |

*Note.* Reward sensitivity was defined as the difference in mean response times between “no reward” and “reward” trials. Positive values for reward sensitivity imply faster responses to “reward” than “no reward” cues. Correlation coefficients shown here are Spearman rhos, significant correlations ($p < .05$ uncorrected) are highlighted in bold. P-values for comparisons of correlations are reported at an uncorrected level and were calculated using Steiger’s approach for dependent overlapping correlations [22] as implemented in the cocor R package [23]. Global IQ was measured using the Wechsler Adult Intelligence Scale (WAIS-IV) [24]. AQ = Autism Spectrum Quotient [8], TAS-20 = Toronto Alexithymia Scale [4,5], BVAQ = Bermond-Vorst Alexithymia Questionnaire [21], BDI-II = Beck Depression Inventory [6,7], IRI = empathy score (sum score excluding “personal distress” subscale) of the Interpersonal Reactivity Index [16,17], SIAS = Social Interaction Anxiety Scale [18], STAI-T = State-Trait Anxiety Inventory – trait [19]. $N = 73$.

**Table S4:** Average task related whole-brain activation in the anticipation phase.

| Anatomical region | Cyto area | Side | Cluster size | MNI coordinates | | | *F* | *p*(FWE) |
|---|---|---|---|---|---|---|---|---|
| | | | | x | y | z | | |
| ***Reward > No reward*** | | | | | | | | |
| Inferior occipital gyrus | hOc1 [V1] | R | 130 | 24 | -97 | -1 | 74.1 | < .001 |
| Middle occipital gyrus | hOc3d [V3d] | L | 212 | -24 | -97 | -1 | 70.0 | < .001 |
| Fusiform gyrus | FG3 | R | 2 | 30 | -55 | -10 | 26.8 | .019 |
| Fusiform gyrus | hOc4v [V4(v)] | L | 3 | -30 | -79 | -13 | 25.4 | .033 |
| Fusiform gyrus | FG3 | L | 1 | -27 | -58 | -13 | 25.1 | .035 |
| ***Social > money*** | | | | | | | | |
| Superior occipital gyrus | hOc2 [V2] | L | 142 | -9 | -97 | 11 | 87.4 | < .001 |
| Cuneus | hOc2 [V2] | R | 103 | 15 | -94 | 11 | 75.3 | < .001 |
| Lingual gyrus | hOc1 [V1] | L | 57 | 0 | -79 | -7 | 58.2 | < .001 |
| Inferior occipital gyrus | hOc4lp | R | 60 | 33 | -85 | -1 | 46.8 | < .001 |
| Fusiform gyrus | FG3 | L | 11 | -30 | -58 | -10 | 41.4 | < .001 |
| Inferior occipital gyrus | hOc4lp | L | 29 | -30 | -88 | -7 | 38.4 | < .001 |
| Fusiform gyrus | FG3 | R | 3 | 30 | -61 | -10 | 27.9 | .014 |

*Note.* Results of contrasts of interest in the anticipation phase, averaged across both groups (ASD and control) and treatments (oxytocin and placebo). P-values are family-wise error (FWE) corrected for whole-brain analyses at the voxel level. The “Cyto Area”-column indicates the cytoarchitectonical area as assigned by the SPM Anatomy toolbox v2.2b [25] if available. Anatomical labels were derived respectively. L = left, R = right. *N* = 71.

**Table S5:** Correlations of reward sensitivity in the ventral striatum and individual difference variables across all participants in the anticipation phase.

| | Left ventral striatum | | | Right ventral striatum | | |
|---|---|---|---|---|---|---|
| | Oxytocin | Placebo | *p*(comparison) | Oxytocin | Placebo | *p*(comparison) |
| ***Money (reward > no reward)*** | | | | | | |
| Age | .008 | .188 | .302 | .151 | .199 | .788 |
| IQ | .035 | -.122 | .372 | .183 | -.149 | .064 |
| AQ | **.249** | -.082 | .056 | **.336** | -.037 | .034 |
| TAS-20 | .198 | -.172 | .034 | .125 | -.097 | .221 |
| BVAQ | -.134 | -.020 | .513 | -.157 | .016 | .338 |
| BDI-II | .157 | -.205 | .037 | .142 | -.203 | .055 |
| IRI | -.124 | .143 | .127 | -.031 | .031 | .732 |
| SIAS | **.383** | -.053 | .010 | **.310** | -.045 | .045 |
| STAI-T | **.244** | .050 | .260 | **.275** | .045 | .193 |
| ***Social (reward > no reward)*** | | | | | | |
| Age | .053 | .055 | .988 | .044 | .039 | .979 |
| IQ | .079 | .136 | .729 | .176 | .107 | .687 |
| AQ | .041 | .087 | .783 | .000 | .052 | .769 |
| TAS-20 | .086 | .145 | .721 | .016 | .210 | .263 |
| BVAQ | -.001 | -.154 | .357 | -.047 | -.179 | .447 |
| BDI-II | -.092 | .111 | .224 | -.138 | .161 | .087 |
| IRI | -.114 | .010 | .458 | -.043 | -.011 | .856 |
| SIAS | -.015 | .089 | .533 | -.033 | .093 | .471 |
| STAI-T | .006 | .062 | .738 | -.073 | .032 | .550 |

*Note.* Reward sensitivity was defined as ventral striatum activation associated with “reward” cues compared to “no reward” cues. Correlation coefficients shown here are Spearman rhos, significant correlations ($p < .05$ uncorrected) are highlighted in bold. P-values for comparisons of correlations are reported at an uncorrected level and were calculated using Steiger’s approach for dependent overlapping correlations [22] as implemented in the cocor R package [23]. Global IQ was measured using the Wechsler Adult Intelligence Scale (WAIS-IV) [24]. AQ = Autism Spectrum Quotient [8], TAS-20 = Toronto Alexithymia Scale [4,5], BVAQ = Bermond-Vorst Alexithymia Questionnaire [21], BDI-II = Beck Depression Inventory [6,7], IRI = empathy score (sum score excluding “personal distress” subscale) of the Interpersonal Reactivity Index [16,17], SIAS = Social Interaction Anxiety Scale [18], STAI-T = State-Trait Anxiety Inventory – trait [19]. $N = 71$.

**Table S6:** Average task related whole-brain activation in the outcome phase.

| Anatomical region | Cyto area | Side | Cluster size | MNI coordinates x | y | z | F | p(FWE) |
|---|---|---|---|---|---|---|---|---|
| ***Reward > no reward*** | | | | | | | | |
| Middle occipital gyrus | hOc4lp | R | 616 | 30 | -94 | 2 | 106.5 | < .001 |
| Fusiform gyrus | FG1 | R | | 33 | -64 | -10 | 36.7 | < .001 |
| Middle occipital gyrus | hOc4lp | L | 749 | -27 | -91 | -4 | 91.9 | < .001 |
| Fusiform gyrus | FG1 | L | | -33 | -64 | -13 | 41.8 | < .001 |
| Middle occipital gyrus | | R | 65 | 30 | -64 | 32 | 33.4 | .002 |
| Angular gyrus | | R | | 27 | -61 | 47 | 30.2 | .005 |
| Superior parietal lobule | 7A (SPL) | L | 34 | -24 | -64 | 53 | 31.4 | .003 |
| ***Social > money*** | | | | | | | | < .001 |
| Lingual gyrus | | R | 3580 | 27 | -49 | -10 | 345.0 | < .001 |
| Fusiform gyrus | FG3 | L | | -27 | -52 | -13 | 295.4 | < .001 |
| Middle occipital gyrus | | R | | 36 | -82 | 14 | 208.4 | < .001 |
| Middle occipital gyrus | hOc2 [V2] | L | 316 | -12 | -103 | 5 | 127.4 | < .001 |
| Cuneus | hOc2 [V2] | R | 213 | 15 | -100 | 11 | 88.7 | < .001 |
| Mid orbital gyrus | Fp2 | R | 265 | 3 | 53 | -13 | 86.5 | < .001 |
| Amygdala | Amygdala (SF) | R | 120 | 21 | -4 | -16 | 72.0 | < .001 |
| Temporal Pole | | R | | 30 | 11 | -28 | 26.0 | .024 |
| Hippocampus | Amygdala (SF) | L | 75 | -18 | -7 | -16 | 71.6 | < .001 |
| Amygdala | | L | | -30 | -4 | -19 | 26.2 | .022 |
| Precuneus | | R | 213 | 3 | -52 | 26 | 55.7 | < .001 |
| WM | | L | | -15 | -46 | 35 | 25.9 | .025 |
| Fusiform gyrus | FG4 | R | 62 | 42 | -46 | -22 | 55.6 | < .001 |
| Superior temporal gyrus | | R | 168 | 48 | -37 | 8 | 44.2 | < .001 |
| Middle temporal gyrus | PGp (IPL) | R | | 57 | -61 | 17 | 33.3 | .002 |
| Superior medial gyrus | | R | 151 | 6 | 56 | 23 | 42.2 | < .001 |
| Superior medial gyrus | | L | | -6 | 56 | 29 | 39.3 | < .001 |
| | hOc1 [V1] | R | 19 | 3 | -79 | -10 | 36.9 | < .001 |
| Superior parietal lobule | 7PC (SPL) | R | 32 | 33 | -46 | 59 | 31.8 | .003 |
| Temporal Pole | | R | 26 | 39 | 20 | -31 | 31.6 | .003 |
| IFG (p. orbitalis) | | R | 9 | 33 | 32 | -16 | 31.4 | .003 |
| Inferior occipital gyrus | hOc4la | R | 3 | 51 | -76 | -1 | 29.4 | .007 |
| Postcentral gyrus | Area 2 | L | 9 | -39 | -34 | 41 | 28.2 | .011 |
| Middle temporal gyrus | | L | 5 | -60 | -4 | -19 | 26.0 | .024 |

**Table S6** (continued).

| Anatomical region | Cyto area | Side | Cluster size | MNI coordinates | | | F | p(FWE) |
|---|---|---|---|---|---|---|---|---|
| | | | | x | y | z | | |
| ***Task x intensity*** | | | | | | | | |
| Lingual gyrus | hOc3v | R | 296 | 24 | -91 | -7 | 85.9 | < .001 |
| Fusiform gyrus | FG1 | R | | 33 | -64 | -10 | 32.0 | .003 |
| Inferior occipital gyrus | hOc4v [V4(v)] | L | 270 | -27 | -88 | -10 | 84.0 | < .001 |
| Angular gyrus | hIP3 (IPS) | R | 45 | 30 | -58 | 47 | 34.6 | .001 |
| Cuneus | hOc3d [V3d] | R | 19 | 9 | -85 | 23 | 29.3 | .007 |
| Fusiform gyrus | FG3 | L | 9 | -33 | -58 | -10 | 27.3 | .015 |

*Note.* Results of contrasts of interest in the outcome phase, averaged across both groups (ASD and control) and treatments (oxytocin and placebo). P-values are family-wise error (FWE) corrected for whole-brain analyses at the voxel level. The “Cyto Area”-column indicates the cytoarchitectonical area as assigned by the SPM Anatomy toolbox v2.2b [25] if available. Anatomical labels were derived respectively. L = left, R = right. *N* = 71.

**Table S7:** Correlations of reward sensitivity of the amygdala and individual difference variables across all participants in the outcome phase.

| | Left amygdala | | | Right amygdala | | |
|---|---|---|---|---|---|---|
| | Oxytocin | Placebo | *p*(comparison) | Oxytocin | Placebo | *p*(comparison) |
| ***Money (reward > no reward)*** | | | | | | |
| Age | -.037 | .078 | .530 | -.111 | .019 | .469 |
| IQ | -.144 | .000 | .433 | -.111 | -.082 | .873 |
| AQ | -.105 | -.118 | .940 | .029 | -.006 | .847 |
| TAS-20 | .143 | -.146 | .115 | .228 | -.009 | .185 |
| BVAQ | .140 | .131 | .964 | .105 | .084 | .910 |
| BDI-II | .146 | -.062 | .259 | .119 | -.021 | .438 |
| IRI | .159 | .039 | .516 | .125 | -.003 | .480 |
| SIAS | .058 | -.073 | .477 | .114 | -.034 | .414 |
| STAI-T | -.028 | -.128 | .585 | .039 | -.067 | .560 |
| ***Social (reward > no reward)*** | | | | | | |
| Age | .126 | .003 | .453 | .024 | -.034 | .751 |
| IQ | -.030 | .221 | .123 | .006 | .107 | .575 |
| AQ | .100 | .026 | .652 | .169 | .004 | .359 |
| TAS-20 | .057 | -.005 | .704 | .153 | .012 | .435 |
| BVAQ | -.092 | -.004 | .595 | -.234 | .022 | .151 |
| BDI-II | .086 | -.103 | .249 | .010 | -.112 | .500 |
| IRI | -.142 | -.220 | .624 | -.078 | -.213 | .448 |
| SIAS | .127 | .081 | .779 | .165 | .174 | .955 |
| STAI-T | .187 | -.055 | .138 | .105 | .104 | .993 |

*Note.* Reward sensitivity was defined as amygdala activation associated with “reward” outcomes compared to “no reward” outcomes. Correlation coefficients shown here are Spearman rhos. No correlation was statistically significant ($p$ < .05 uncorrected). P-values for comparisons of correlations are reported at an uncorrected level and were calculated using Steiger’s approach for dependent overlapping correlations [22] as implemented in the cocor R package [23]. Global IQ was measured using the Wechsler Adult Intelligence Scale (WAIS-IV) [24]. AQ = Autism Spectrum Quotient [8], TAS-20 = Toronto Alexithymia Scale [4,5], BVAQ = Bermond-Vorst Alexithymia Questionnaire [21], BDI-II = Beck Depression Inventory [6,7], IRI = empathy score (sum score excluding “personal distress” subscale) of the Interpersonal Reactivity Index [16,17], SIAS = Social Interaction Anxiety Scale [18], STAI-T = State-Trait Anxiety Inventory – trait [19]. $N$ = 71.
